# Barriers and Facilitators of Expedited Partner Therapy in U.S. Emergency Departments: A Nationwide Qualitative Study

**DOI:** 10.1101/2025.09.16.25335878

**Authors:** Rachel E Solnick, Rafael Cortes, Ethan Chang, Paul Dudas, Alan Cetkovic, Ricardo Londoño-Barreras, Maaz Munawar, Carlin Pendell, Aisling Shi Zhao, Keith E. Kocher, Roland C. Merchant

**Affiliations:** Icahn School of Medicine at Mount Sinai, NY, New York; Previously at: National Clinical Scholars Program, Department of Emergency Medicine, University of Michigan, Ann Arbor, Michigan, USA; University of Southern California, Los Angeles, California, USA; Previously at: University of Michigan, Ann Arbor, Michigan, USA; SUNY Downstate College of Medicine, New York, New York, USA; Department of Psychology and Neuroscience, University of Colorado, Boulder, Boulder, Colorado, USA; Department of Family Medicine, International Community Health Services, Seattle, Washington, USA; Department of Emergency Medicine and Learning Health Sciences, University of Michigan, Ann Arbor, Michigan, USA; Department of Emergency Medicine, Morsani College of Medicine at University of South Florida, Tampa, Florida, USA

**Keywords:** Sexually Transmitted Diseases, Sexually Transmitted Infection, Partner therapy, implementation science, expedited partner therapy

## Abstract

**Background:** Expedited partner therapy (EPT) is recommended for sex partners who might not seek care otherwise. Despite its high relevance in the management of sexually transmitted infections (STIs), EPT has had limited uptake in emergency departments (ED). We assessed barriers to and facilitators that could assist in increasing use of EPT in the EDs.

**Methods:** We conducted semi-structured interviews of 18 ED medical directors and key medical personnel from institutions that offer or were interested in offering EPT, as identified through our prior research. We developed and piloted an interview guide based on the Consolidated Framework for Implementation Research (CFIR). Interviews were recorded, transcribed, and analyzed using a thematic analysis. Themes were developed through iterative coding and team discussion.

**Results:** Participant-reported barriers to implementing EPT included perceived legal liability, unclear clinical workflows, electronic health record (EHR) challenges when prescribing for a non-patient, ambiguity regarding duty to the partner, concerns about patient safety, and sexual health stigma. Facilitators included defined task-sharing, streamlined EHR processes, and leadership support. Participants emphasized the ED’s potential to expand access to care for high-risk populations, provided that EPT implementation did not disrupt clinical operations.

**Conclusions:** Understanding barriers and facilitators to EPT in EDs can inform the development of effective implementation strategies. While non-traditional prescribing processes pose challenges to routine adoption, experiences from EDs with established protocols suggest that these obstacles can be overcome.

## INTRODUCTION

Cases of new sexually transmitted infections (STIs) remain high in the United States (US). Over 2.4 million cases of chlamydia, gonorrhea, and syphilis were reported in 2023, according to the Centers for Disease Control and Prevention (CDC).^1^ Emergency departments (EDs) play a critical role in STI care, particularly for patients who identify the ED as a usual source of care.^2^ In fact, ED visits for STIs in the US are continuing to rise and frequently yield higher STI test positivity rates than outpatient settings. ^3,4^

Traditional approaches to treating sex partners for STIs remain inadequate. Patients notifying their own partner is the most commonly used way sex partners are informed of their need for treatment, but this method consistently results in low treatment rates.^5,6^ CDC surveillance data indicate that partner notification rates range from 29% to 59%^7^, and only about 35% of partners receive treatment.^8^

Expedited Partner Therapy (EPT) allows clinicians to treat the sex partners of patients with certain STIs without a clinical exam. EPT reduces reinfection by up to 29%^9^ and has been recommended by the CDC since 2006. However, uptake in EDs—a key setting for STI care—remains low.^10^ Patients seen in EDs or hospitals are significantly less likely to be offered EPT than those seen in STI clinics,^11^ and only 9% of ED clinicians in a New York City study reported using it.^12^

Previous studies on EPT implementation have focused on pediatric ED populations or excluded EDs altogether.^13–15^ Given the complex, individual and system-level challenges of integrating EPT into ED workflows, we applied an implementation science framework to better understand real-world barriers to and facilitators that could assist in increasing use of EPT in EDs We aimed to identify obstacles at EDs interested in EPT but not yet implementing it, and also to characterize the strategies that enabled successful adoption at EDs with active EPT programs.

## METHODS

Between July 2020 and February 2021, we conducted semi-structured interviews of ED medical directors and key medical personnel from institutions that offer or are interested in offering EPT. This study was deemed exempt by the University of Michigan Institutional Review Board (IRB HUM00196451). All participants provided informed consent prior to participation. We report our methods in accordance with the Consolidated Criteria for Reporting Qualitative Research (COREQ) checklist. ^16^

### Recruitment and Purposeful Sampling

For this project we first recruited medical directors from institutions that had implemented or whose directors had expressed interest in implementing EPT, based on responses to our previously published nationwide survey.^10^ With the assistance of the medical directors and administrative leaders who agreed to participate, we employed snowball sampling to identify additional key medical personnel (nurses, Emergency medicine (EM) resident physicians, program directors, EM physician, and pharmacists) from those institutions who also could provide insight into EPT processes. Participants were selected based on their involvement or knowledge of EPT at their institutions. We contacted 20 medical directors and key informants via email; 18 responded and agreed to participate in an interview.

### Instrument Development

We developed a semi-structured interview guide informed by the Consolidated Framework for Implementation Research (CFIR) to explore multilevel barriers and facilitators to EPT implementation in emergency departments. The guide drew on prior EPT literature^17,18^ and key CFIR domains, including intervention characteristics, outer and inner setting, individual characteristics, and process.^19^ It was organized into five sections and reviewed by an implementation expert, then pilot-tested with three unaffiliated emergency clinicians. Feedback led to minor wording changes. The final guide included 14 core questions with optional probes mapped to CFIR constructs.

### Data Collection

We conducted semi-structured, open-ended virtual interviews with participants. Interview questions focused on experiences with, awareness of, and opinions on EPT, as well as perceived barriers and facilitators to its implementation in the ED. The primary investigator (RES, EM physician with formal training in qualitative methods) conducted the interviews alongside three study team members (research assistants with training in qualitative methods) to ensure comprehensive data capture. Participants were aware of the interviewer’s role as a clinician-researcher and the purpose of the study. The interviewer’s role may have facilitated rapport, but it also posed a potential bias, which was addressed through team-based reflexivity during analysis. No repeat interviews were conducted. Interviews were transcribed verbatim by a professional transcription service. Transcripts were not returned to participants for their review. A total of 18 interviews were conducted, with no refusals after invitations, each lasting approximately 20–50 minutes. Interviews continued until thematic saturation was reached. All participants received a gift card after completing the interview.

### Analysis

We used a modified version of Braun and Clarke’s thematic analysis method:^20^ (1) familiarization with the data through repeated reading of transcripts, (2) generating initial codes using both inductive and Consolidated Framework for Implementation Research (CFIR)^19^-informed deductive strategies, (3) searching for themes by grouping related codes into broader patterns, (4) reviewing themes for coherence and distinctiveness, (5) defining themes based on their organizing concepts, and (6) producing the narrative synthesis with illustrative quotations. Coding and theme refinement were conducted iteratively by the research team through regular debriefing meetings, with consensus reached on all final themes. Each interview was coded by at least two researchers to ensure reliability, with no major disagreements. The data were analyzed using NVivo 10.0 software.

## RESULTS

### Data collected

We interviewed 18 participants from 15 US EDs, including 11 medical directors or administrative leaders, two pharmacists, one director of pediatric ED, one follow-up nurse, one EM physician, one EM resident physician, and one EM program director (Table 1 for participant characteristics).

**Table 1.** Descriptive Summary of Participants and Emergency Departments.

|  |  |  |
| --- | --- | --- |
| **Participant Characteristic (n=18 participants)** | **Number** | **Percentage or Range** |
| **Role** |  |  |
| Medical Director/ Administrative leader | 11 | 61% |
| Pharmacist | 2 | 11% |
| Director of Pediatric EM | 1 | 6% |
| Follow-up Nurse | 1 | 6% |
| Faculty member | 1 | 6% |
| Resident | 1 | 6% |
| Program director | 1 | 6% |
| **Sex** |  |  |
| Male | 14 | 78% |
| Female | 4 | 22% |
| **ED Characteristics (n=15 EDs)** |  |  |
| EPT Implementer | 6 | 40% |
| Departmental-level policy | 4 | 27% |
| Individual EPT prescriber | 2 | 13% |
| **Medicaid, median% (IQR)** | 36% | (23%-43%) |
| **ED Volume** | 64,875 | (62,541—83,728) |
| **Region** |  |  |
| Northeast | 4 | 27% |
| Midwest | 6 | 40% |
| South | 2 | 13% |
| West | 3 | 20% |
| *Note: Administrative leaders included Chief of Emergency Services, Director of Operations for Emergency Department, Vice Chair of Clinical* |  |  |

|  |  |  |
| --- | --- | --- |
| Participant Characteristic (n=18 participants) | Number | Percentage or Range |
| Role |  |  |
| Operations |  |  |

Of the 15 sites, around 40% had implemented EPT at least partially. Participants from two of the sites characterized its implementation as an individual approach rather than a policy-based approach at their institutions. Participants from EDs that had not formally adopted EPT expressed interest in adopting it at their EDs.

The interviews revealed common challenges across institutions and informed our summarization of barriers and facilitators influencing EPT implementation in EDs, which we present under the following thematic categories.

### ED Is a Critical but Constrained Setting for Sexual Health Care

Several health care professionals viewed STI care as outside the traditional EM scope of acute conditions, but they recognized the ED’s role in compensating for gaps in primary and sexual health care. STI management was considered more appropriate for outpatient clinics due to the need for follow-up, HIV testing, and continuity of care. However, EDs were often noted as the only point of access for many underserved patients due to systemic limitations in healthcare.

> *“*…*because we have such a fragmented healthcare system, it’s our obligation to provide that safety net in the emergency departments*.*”* (EPT009 Medical Director)

Local contexts influenced how participants perceived their responsibilities in delivering STI services in the ED. A few participants noted that at their sites, there were well-developed primary care networks or STI clinics, which resulted in less pressure to provide comprehensive sexual health services. While another participant noted that urgent care services encompassed lower-acuity patients, resulting in less STI care delivered in the ED. On the other hand, sites with high frequencies of STI complaints, and community organizations recognized EPT as a local priority and developed measures to offer it as part of a reproductive health program.

Finally, for some, the COVID-19 pandemic specifically highlighted the importance of public health within EM:

> *“As COVID has happened, we understand that we [emergency medicine] have a role in public health. And that that’s being more embraced than it was in the past*.*”* (EPT006 Medical Director)

### Normalizing Sexual Health in the ED: Overcoming Cultural Discomfort and Systemic Friction

### Discomfort discussing sexual health

The majority of participants expressed discomfort discussing sexual health with patients, especially in contexts deviating from cisgender or heterosexual sexual practices. Some felt unequipped to navigate conversations around sex, while others noted that patients’ own stigma also interfered with effective care, as described by the resident.

However, to reduce awkwardness, some providers adopted a more positive framing. For example, one described encouraging patients, especially women, to feel empowered about their sexuality:

> *“I’ve been playing with a spiel where I’ve really encouraged people, particularly my female patients, to feel empowered about their sexuality…*.. *emphasize that it’s really about them taking control and being able to have as many partners as they want by just taking a few precautions, and it’s very well received*.*”* (EPT004 Medical Director)

### Low awareness of EPT clinical pathways

Several participants described low awareness of EPT, with three not realizing EPT was offered at their institutions (EPT007 Medical Director, EPT009 Medical Director, EPT003 Program Director). One participant described how this knowledge void among faculty created a barrier among residents to prescribe EPT, who would otherwise have been early adopters:

> *“Part of the problem is that I don’t think most of us even knew that [EPT] was an option because it had not been disseminated to the faculty… The residents often are drivers of culture. I’ve maybe been asked by one resident whether we could do partner therapy, and I was like, “I don’t think that’s legal*.*”“* (EPT003 Program Director)

One pharmacist shared similar concerns about decreased awareness, describing how, even though helping achieve a protocol it got lost over time:

> *“*…*it totally got lost in the shuffle… but then [ED staff and providers] just kind of forgot about it”* (EPT012 Medical Director).

Providers discussed that efficiency and clarity were key to implementing a successful protocol in the ED, making a streamlined, friendly workflow in a fast-paced environment like the ED. One participant noted:

> *“If we could streamline a process, I think it’s something my group would have no trouble with*… *If there was one button you could press,’EPT done, here’s your paper script,’ I believe people would be happy to do it*.*”* (EPT006 Medical Director)

Several participants expressed that team-based care was a facilitator. One noted that at their site, the ED pharmacists play an active role in the EPT implementation. Patients could easily stop by the ED’s 24/7 pharmacy to retrieve medications for themselves and their partners (EPT011 Medical Director).

### Perceived risks and system constraints complicate non-traditional STI care models

Because the nature of EPT requires the prescription of medication to a person with whom the provider does not have a traditional physician-patient relationship, the main barriers derive from this ambiguity of fiduciary responsibility to the partner in terms of legal protection, patient safety, and the design of the EHR.

### Medicolegal and safety concerns

A frequent barrier referenced by most participants was medico-legal concerns. Several participants expressed concerns about potential litigious consequences, despite being aware of the existing legal protections.

> *“Just because there’s a law in place doesn’t mean you’re protected from an adverse outcome…”* (EPT003 Medical Director).

One medical director who was willing to perform EPT was aware that other physicians would be less willing to use EPT due to the increased liability this practice carries (EPT002 Medical Director). Another provider noted the added burden of documenting discussions with patients to mitigate liability (EPT003 Medical Director).

A few of the interviewees mentioned safety concerns related to EPT. One was worried about the possibility of partner violence when the partner is notified. (EPT004 Medical Director). While others were worried about the diagnosis uncertainty and potential overtreatment of people, and possible potential allergies the partner might have.

### EHR challenges

Many participants stated that their institutions lacked the necessary EHR functionality to correctly implement EPT, especially if it was for an individual without a pre existing record in the EHR. One participant noted that even institutions with EPT protocols struggle due to the different EHR systems in place across affiliated hospitals. While others indicated that, due to state laws, there is an EHR restriction that requires patients’ names and addresses to fill prescriptions:

> *“*…*Without a visit, you can’t generate the prescription… How do you make sure that you’re getting the right information to the right pharmacy?”* (EPT015 Medical Director)

In contrast to these issues, at sites that successfully implemented EPT, the EHR facilitated prescribing by standardizing the EPT process across multiple hospitals.. Another director explained how using the EHR improved what was previously an onerous paper-based process that was met with a lot of staff resistance to one that was more easily adopted across sites:

> *“*…*the nice thing was that it was fairly self-explanatory, didn’t require a lot of training for providers and so it was easy to promote… the fact that it is essentially kind of a two clicks, a signature, and you’ve kind of done the right thing helped with that rollout and adoption*.*”*(EPT005 Medical Director)

### Champions Enable Change, but Influence Is Shaped by Role and Scope

Participants from sites that successfully implemented EPT often mentioned champions who leveraged authority and influence in their leadership roles and had the commitment and capacity to follow through on EPT implementation. Sites that were interested in EPT, but did not actively use it, either lacked champions or had organizational hierarchies that hindered process change.

#### Leadership

Some medical directors had led EPT efforts through policy changes or protocol development. For example, one medical director partnered with pharmacists through a collaborative practice agreement, allowing them to prescribe EPT independently.

> *“*… *having a leadership role in the department, and in the system, is helpful in that we can champion these really important causes about* … *public health, and get people to listen and understand*.*”* (EPT006 Medical Director)

However, not all directors had the same capacity to drive change. Some reported that institutional hierarchies had constrained their ability to implement new practices.

#### Influence

Beyond being in positions of authority, influence also facilitated change. Senior physicians were able to align EPT efforts with local community needs and shape practice norms among peers. In one case, providers responded to advocacy from the YWCA, a local organization serving sexual assault victims, and aligned ED services accordingly.

#### Longitudinal Commitment

Long-term commitment was also essential. Participants expressed concern that resident-driven initiatives often failed to persist due to turnover. Without a stable, invested advocate embedded within the department, EPT programs were likely to be forgotten over time. Moreover, medical directors often had competing demands, such as meeting public metrics, which made it difficult to champion initiatives like EPT without external incentives or institutional support.

> *“Medical directors are terrible champions often… They will get passionate about door-to-needle time. That’s because they don’t want to look bad nationally*.*”* (EPT003 Medical Director)

## DISCUSSION

This study of ED medical directors and key medical personnel in EDs at institutions that had implemented or whose directors had expressed interest in implementing EPT reveals a strong enthusiasm for addressing sexual and reproductive health public health needs. However, our findings confirm that many barriers to implementing EPT in US EDs exist at individual, institutional, and system levels, many of which have been previously documented.^21,22^

A key barrier is the clinician’s reluctance to prescribe EPT due to medico-legal concerns, even in states where the practice is legally protected. This hesitancy often results from a lack of awareness regarding EPT laws.^23^ Providers worry about the potential for overtreatment, especially in the absence of a direct clinical relationship with the partner. Prior studies have shown that such uncertainty can contribute both to inappropriate antibiotic use and to missed treatment opportunities—especially for chlamydia and gonorrhea, where women are less likely than men to receive appropriate therapy.^24^

Institutional barriers also limit EPT uptake, including competing priorities, limited time, and EHR systems not configured to support prescribing.<u>^25^</u> The absence of streamlined order sets adds friction for clinicians. These workflow challenges, along with provider uncertainty and limited infrastructure, have been linked to low EPT adoption in EDs.^26–28^ Addressing these challenges may require targeted EHR modifications, leadership engagement, and workflow integration to support sustainable use of EPT in the ED setting.

Some EDs successfully implemented EPT through localized, flexible strategies. These strategies include staff-driven policy changes, delegation to non-physician personnel, and EHR adaptations that enable proper documentation and prescribing. This success was more common in high-need areas or where institutional and community support was strong, underscoring the potential of targeted, context-specific approaches.

Healthcare professionals’ discomfort discussing sexual health emerged as another major obstacle. Participants reported challenges initiating conversations with patients about STIs. Our findings reinforce that persistent communication barriers and stigma may limit not only patient counseling but also the routine use of EPT.^29–31^ Stigma surrounding sexual health discussions, especially with women and adolescents, remains a common barrier.^29,30^ A 2016 study recommended using nonjudgmental language and creating supportive environments for sexual health discussions with patients.^31^

Low awareness of EPT and disconnects with pharmacy workflows remain key implementation barriers. Across sites, participants reported limited awareness of EPT laws and procedures, with some unaware that their own institution offered EPT. This lack of knowledge extended to both attending physicians and residents, constraining adoption and creating confusion around legality. These findings align with prior studies showing poor provider awareness, including one where over two-thirds of pediatric ED clinicians were unaware EPT was legal.^23^In our study, knowledge gaps also contributed to a disconnect between ED prescribers and pharmacy workflows. Participants described difficulty ensuring prescriptions were filled, especially when partners’ names were omitted, an issue previously linked to high pharmacy rejection rates.^33,34,35^ While some sites had success with integrated ED pharmacy teams and EHR-supported workflows, others struggled with fragmented systems and legal requirements that hampered prescribing. These challenges were more pronounced in areas without strong community partnerships or institutional support, reflecting wider variability in EPT implementation across the country.^12,21,36^

In summary, the results from this study reinforces and expands on prior research by demonstrating that EPT implementation in EDs is hindered by an interplay of legal ambiguity, system limitations, stigma, knowledge gaps among providers and pharmacists. Overcoming these challenges will require multifaceted solutions, including policy reforms to clarify and support the use of EPT, education for clinicians and pharmacists, improvements to EHRs, and deliberate efforts to normalize conversations about sexual health.

## LIMITATIONS

This study has several limitations. Most participants were medical directors, which may have introduced selection bias and narrowed the range of perspectives. However, purposeful sampling of high-interest EDs and a 69% survey response rate support the robustness of our sample. Findings may not generalize to community EDs, as all interviews were conducted in academic settings to better understand implementation in educational environments, thereby excluding community EDs. Finally, the interviews took place during the COVID-19 pandemic, which likely affected directors’ capacity to prioritize new initiatives like EPT.

## CONCLUSION

Overcoming key barriers, particularly medico-legal concerns and the lack of streamlined EHR workflows, is crucial to expanding EPT implementation in US EDs. Although not the ideal venue for managing sexual health care, the ED plays a vital role due to gaps in primary care access. Directors supported EPT’s public health mission but were wary of its complexity and risks. Further research should develop and test implementation strategies tailored to diverse ED environments.

## Data Availability

De-identified excerpts supporting the findings are included in the manuscript and its supplementary materials. Full interview transcripts are not publicly available to protect participant confidentiality.

## Author contributions

Rachel E. Solnick: Conceptualization, Methodology, Validation, Formal analysis, Investigation, Writing – Original Draft, Data Curation.

Rafael Cortes: Validation, Formal analysis, Investigation, Writing – Original Draft

Ethan Chang: Formal analysis, Investigation, Writing – Original Draft

Paul Dudas: Formal analysis, Investigation

Alan Cetkovic: Formal analysis, Investigation

Ricardo Londoño-Barreras: Writing – Original Draft, Writing – Review & Editing

Maaz Munawar: Formal analysis, Investigation, Writing – Original Draft

Carlin Pendell: Formal analysis, Investigation, Writing – Original Draft

Aisling Shi Zhao: Methodology, Validation, Formal analysis, Investigation, Writing – Original Draft

Keith E. Kocher: Conceptualization, Methodology, Supervision, Writing – Review & Editing

Roland C. Merchant: Methodology, Supervision, Writing – Review & Editing

## Statements and declarations

De-identified excerpts supporting the findings are included in this article and its supplementary materials. Full interview transcripts are not publicly available to protect participant confidentiality.

## Ethical considerations

This study was deemed exempt by the University of Michigan Institutional Review Board (IRB HUM00196451). All participants provided informed consent. No patients were involved, and all data were de-identified prior to analysis.

## Funding statement

Dr. Solnick was supported by the Institute for Healthcare Policy and Innovation at the University of Michigan National Clinician Scholars Program and by grant K23MH136923-01 from the National Institutes of Mental Health during this work.

**Supplemental Table.**
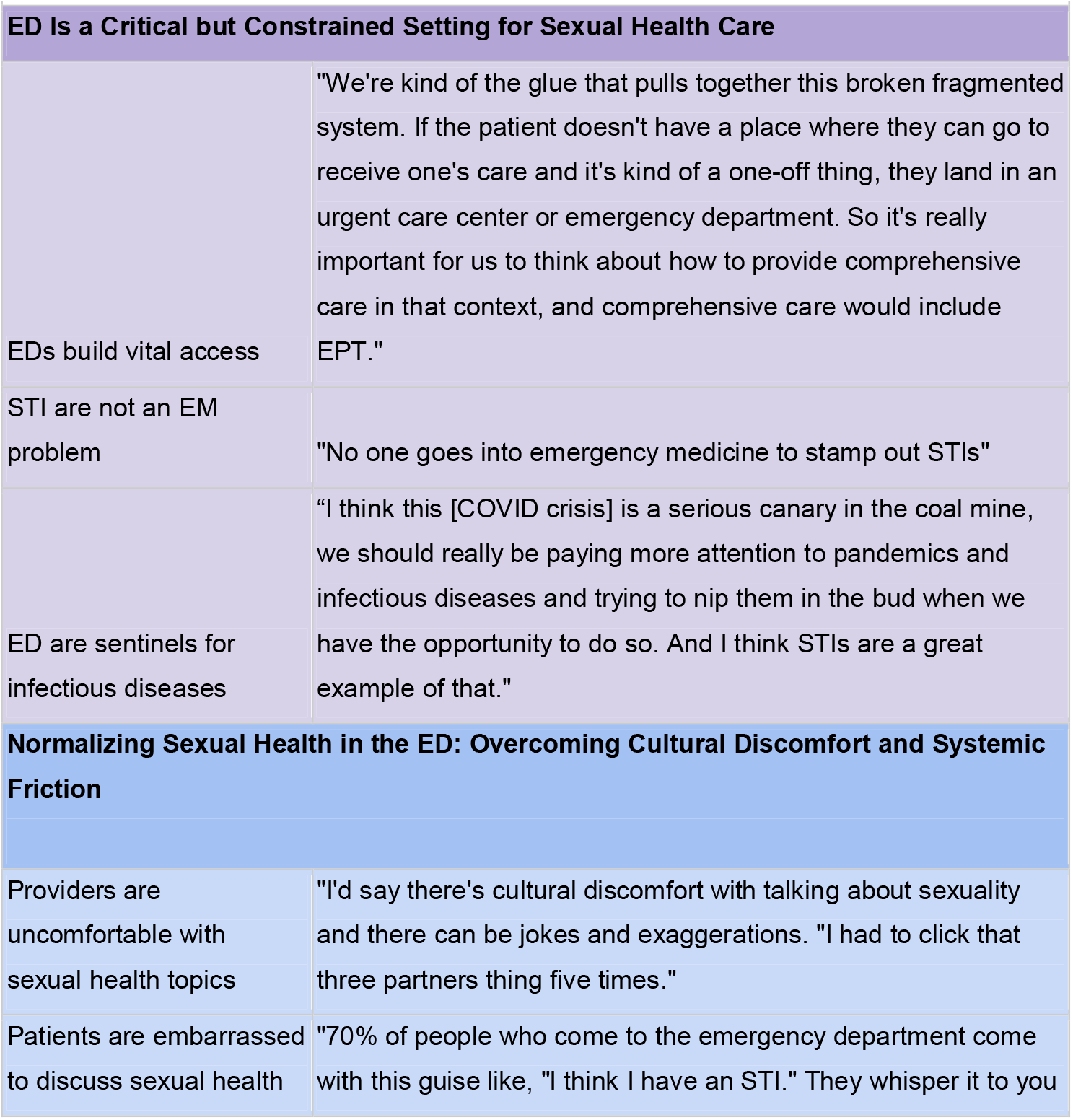
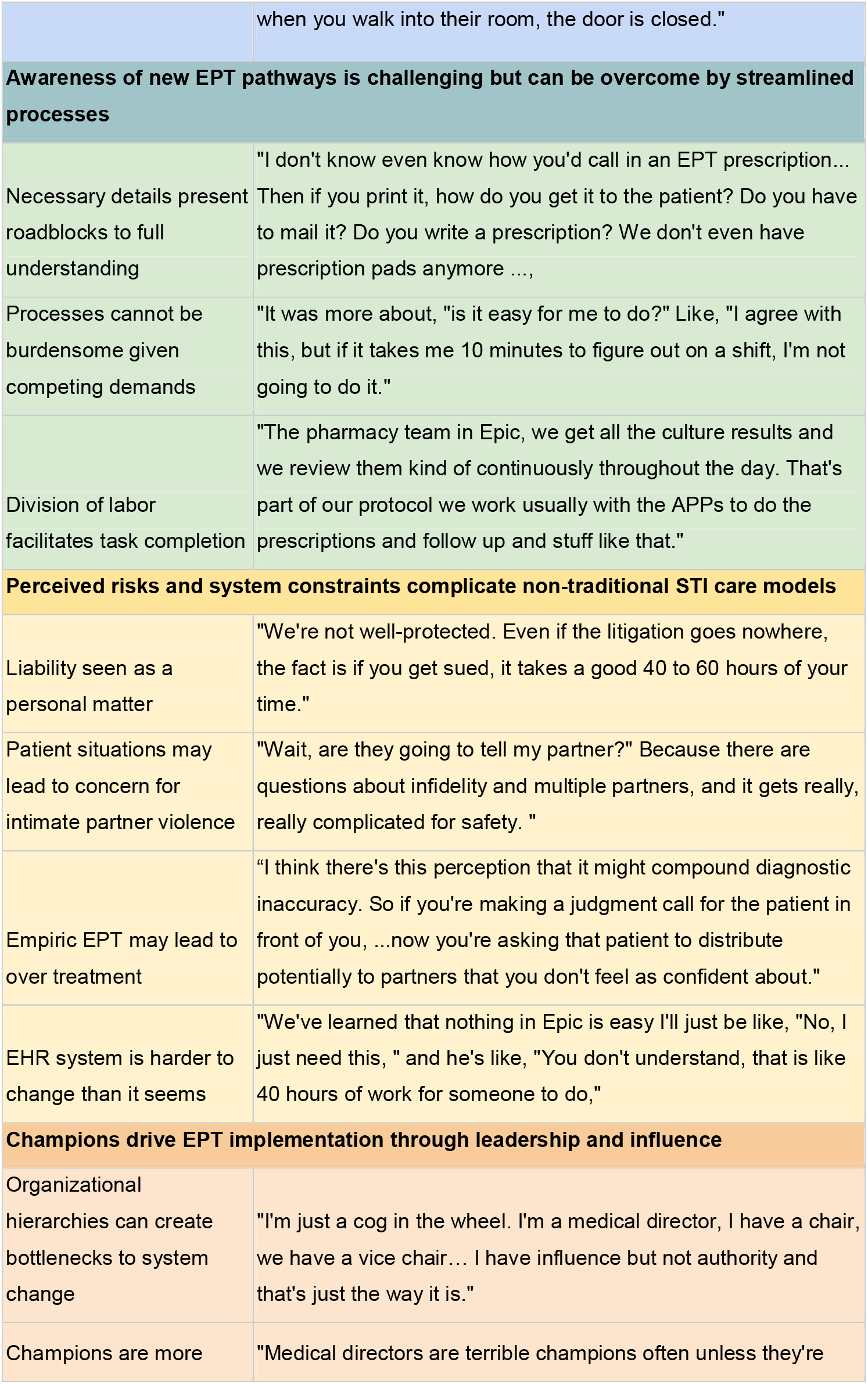
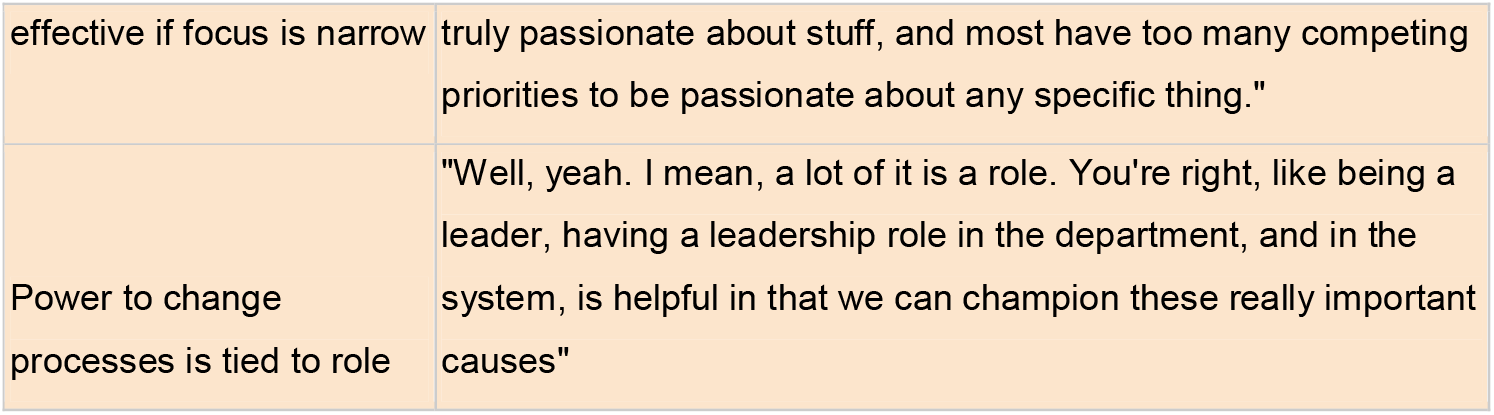
Themes and Supporting Quotes Related to Key Informants and Medical Directors’ Views on ED EPT.

